# Characterization and Mitigation of Model Bias in Parametric Mapping of Dopamine Response to Behavioral Challenge

**DOI:** 10.1101/2021.06.29.21259715

**Authors:** Michael A. Levine, Joseph B. Mandeville, Finnegan Calabro, David Izquierdo-Garcia, Julie C. Price, Beatriz Luna, Ciprian Catana

## Abstract

Compartmental modeling of ^11^C-raclopride (RAC) is commonly used to measure dopamine response to intra-scan behavioral tasks. Bias in estimates of binding potential (BP_ND_) and its dynamic changes (ΔBP_ND_) can arise when the selected compartmental model deviates from the underlying biology. In this work, we characterize the bias associated with assuming a single target compartment and propose a model for reducing this bias by selectively discounting the contribution of the initial uptake period.

**Methods:** 69 healthy young adult participants were scanned using RAC PET/MR while simultaneously performing a rewarded behavioral task. BP_ND_ and ΔBP_ND_ were estimated using an extension of the Multilinear Reference Tissue Model (MRTM2) with the task challenge encoded as a Heaviside step function. Bias was estimated using simulations designed to match the acquired data and was reduced by introducing a new model (DE-MRTM2) that reduces the biasing influence of the initial uptake period in the modeled estimation of BP_ND_ for both simulations and participant data.

**Results:** Bias in ΔBP_ND_ was observed to vary both spatially with BP_ND_ and with the assumed value of k_4_. At the most likely value of k_4_ (0.13 min^-1^), the average bias and the maximum voxel bias magnitude in the nucleus accumbens were estimated to be 1.2% and 3.9% respectively. Simulations estimated that debiasing the contribution of the first 27 minutes of acquired data reduced average bias and maximum voxel bias in the nucleus accumbens ΔBP_ND_ to -0.3% and 2.4% respectively. In the acquired participant data, DE-MRTM2 produced modest changes in the experimental estimates of striatal ΔBP_ND_, while extrastriatal bias patterns were greatly reduced. DE-MRTM2 also considerably reduced the dependence of ΔBP_ND_ upon the first-pass selection of k_2_’.

**Conclusion:** Selectively discounting the contribution of the initial uptake period can help mitigate BP_ND_- and k_4_-dependent bias in single compartment models of ΔBP_ND_, while also reducing the dependence of ΔBP_ND_ on the first-pass estimation of k_2_’.

## Introduction

Dopamine responses to experimental interventions—both pharmacological^1-3^ and behavioral^4, 5^—are frequently quantified by measuring changes in non-displaceable binding potential (BP_ND_) using ^11^C-raclopride (RAC) Positron Emission Tomography (PET) tracer kinetic modeling. Depending on the design of the study, the RAC kinetic modeling may either employ a two-tissue compartment model configuration and sampled arterial blood as an input function or it may make simplifying assumptions in order to apply a one tissue compartment model configuration and/or derive the input function from a reference region devoid of specific binding^6, 7^.

When the assumptions underlying the models used to analyze RAC are violated, bias is introduced into the modeled estimates of BP_ND_ and its change over time (ΔBP_ND_). In the context of experimental RAC challenge studies, this bias can be difficult to disentangle from true responses, especially behavioral responses^4^, which tend to be much smaller than the responses to pharmacological challenges^8^ that are typically used to probe dopaminergic function^9^.

Specifically, one common source of model bias arises from fitting a model with a one-tissue compartment configuration to data that is more accurately represented by two-tissue compartments^10, 11^. Single-tissue compartment configurations are often employed to improve model stability and convergence, rendering such bias a necessary tradeoff. This bias may be compounded if the model is further reduced by fixing the rate of efflux from the reference region (k_2_’) to constrain the parameter space and thereby facilitate robust parametric mapping^12, 13^. These biases can be reduced without increasing the number of fit parameters by adjusting the model to better fulfill its simplifying assumptions. In this work, we consider the one-tissue model assumption of fast-exchange between the nondisplaceable and specifically bound compartments. As this assumption may be unsafe before steady-state is established, we propose a new model: the Debiased Extended Multilinear Reference Tissue Model (DE-MRTM2). DE-MRTM2 eliminates the contribution of the initial uptake period to the estimation of BP_ND_ and ΔBP_ND,_ thereby reducing model-based bias in estimates of behavioral task response.

First, we performed simulations to replicate the conditions of the study, such that model bias could be estimated relative to a known ground truth. The parameters of the simulation were selected based on analysis of a very large human cohort subjected to a within-scan reward task, plus a second RAC dataset that included arterial blood plasma samples. From these simulations, the extent of the model bias in the rewarded task study was estimated and a method for reducing the bias attributable to the initial uptake period was evaluated. Finally, we performed kinetic analyses of this large human rewarded-task RAC study using one-compartment methodologies with and without simulation-motivated bias reduction.

## Methods

### Participants and Data Acquisition

This retrospective analysis focused on a set of healthy adult participants (N = 69, age 18-30 years) who were scanned using simultaneous RAC PET/Magnetic Resonance (MR) with a rewarded task challenge paradigm. The data was acquired at the University of Pittsburgh Medical Center on a Siemens Biograph mMR scanner (Erlangen, Germany). All participants gave written informed consent and the study was approved by the local Institutional Review Board. All participants were imaged in headfirst supine position.

A bolus of RAC (661-802 MBq) was delivered at the start of the scan followed by continuous infusion (KBol = 105 min), and PET data were acquired in listmode format for 90 minutes. Attenuation maps were generated from a T1-weighted anatomical MR sequence (MPRAGE) using a pseudo-CT method^14^. PET data were iteratively reconstructed using uniform 3-minute frames (OP-OSEM, 3 iterations, 21 subsets, no filter). Frame-based registration for motion correction was performed using the volume closest to the start of the reward task (t = 40 min) as the reference.

The simultaneously acquired MR focused primarily on fMRI sequences (2 resting state, 6 task). Also included were routinely acquired head MRI sequences (MPRAGE, T1 mapping, localizer, gradient-echo field mapping, ultrashort TE, TurboFLASH, and magnetization transfer ratio). Further information about the data set can be found in a previous report^15^.

### Simulation Design

To distinguish bias effects from genuine task response, simulated PET data were generated without a behavioral challenge to examine the impact of model bias alone. Simulated PET time activity curves were generated for every voxel in the brain using a two-tissue compartment model configuration. Fully simulating this set of four-dimensional timeseries PET data required an arterial input function (C_P_) and values of K_1_, k_2_, k_3_, and k_4_ for every voxel in the brain (**Figure 1**).

**Figure 1:**
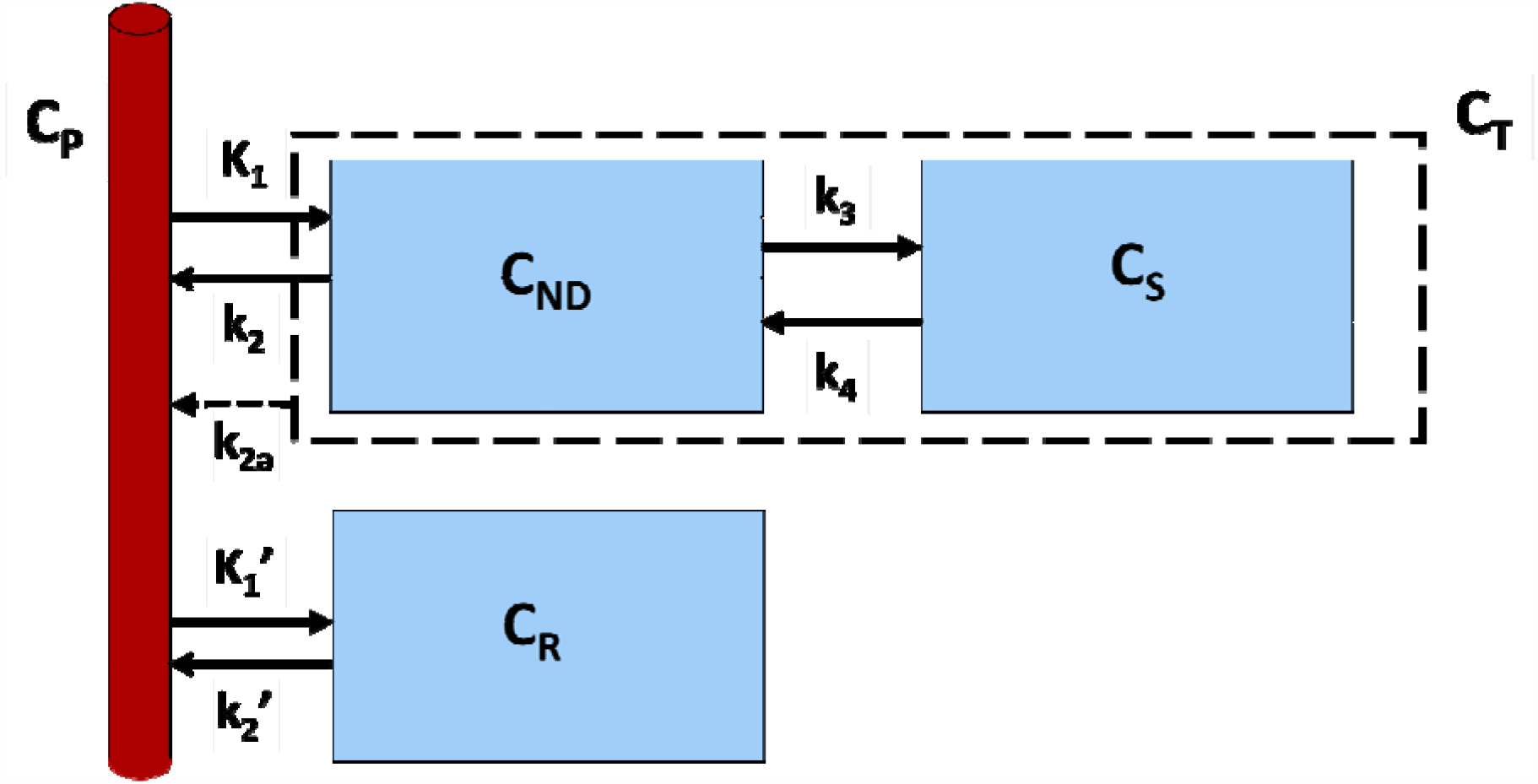
Compartmental model representing PET data. The target tissue (C_T_) is represented by two compartments: the non-displaceable compartment (C_ND_) and the specifically bound compartment (C_S_). The reference tissue (C_R_) is represented by a single compartment and has no specific binding. When C_T_ is simplified to a one-tissue model (dashed lines), the effects of k_2_, k_3_, and k_4_ (and therefore BP_ND_) are collected in a single parameter: the apparent efflux coefficient (k_2a_).

To maximize the salience of the simulation experiments, these input parameters to the simulations were chosen to match the acquired data as closely as possible. Individual values of K_1_, k_2_, and k_3_ for each voxel were generated using the definitions of their associated macroparameters: R_1_ and BP_ND_ (**Eqs. 1-3**)^7^.

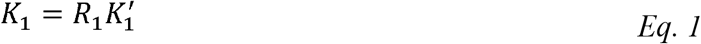

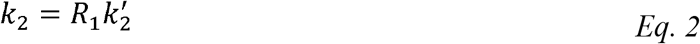

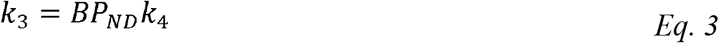

To determine the most appropriate values of R_1_ and BP_ND_, voxelwise analysis with MRTM2^13^ was performed on the acquired PET data and maps of R_1_ and BP_ND_ were averaged across all participants to serve as “ground truth” maps for input to the simulations (**Figure 2**).

**Figure 2:**
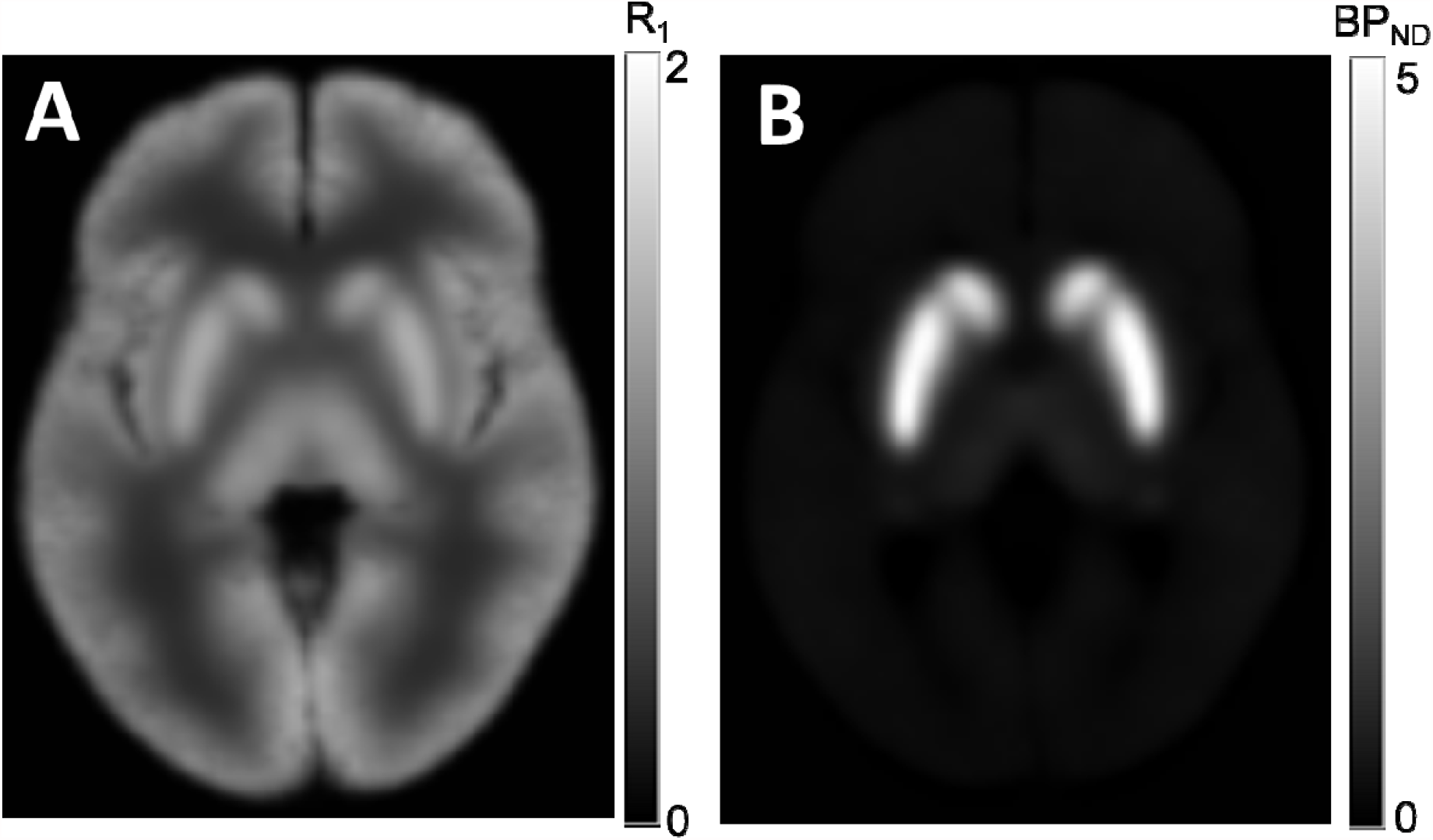
Ground truth maps used as inputs for simulations. (A) Voxelwise R_1_ fit by MRTM2 averaged across all participants. (B) Voxelwise BP_ND_ fit by MRTM2 averaged across all participants.

To estimate the rate constants between blood plasma and the reference tissue compartment (K_1_’, k_2_’), we obtained a control data set, which also administered RAC to healthy adult volunteers, while additionally sampling arterial blood^16^. The reference region (cerebellum) TAC was fit to the measured blood plasma TAC using a nonlinear one tissue compartmental model to estimate K_1_’ and k_2_’. The average estimated values across four participants (K_1_’ = 0.22 mL cm^-3^ min^-1^, k_2_’ = 0.58 min^-1^) were selected for use in the simulations.

To estimate the arterial input function (C_P_) for the task-group subjects, a family of C_P_ curves was used with the chosen values of K_1_’ and k_2_’ to simulate cerebellar TACs. The simulated TAC which best fit the average cerebellum TAC across study participants was selected as the most representative and the C_P_ curve used to generate that TAC was used as the basis for further simulations.

In contrast to the selection of the other compartmental microparameters, a robust means of assigning voxelwise estimates to k_4_ was not available. Therefore, individual simulations fixed k_4_ to a single value within a physiologically plausible range (k_4_ = 0.07, 0.13 min^-1^)^11, 17^ in order to calculate levels of model-induced bias.

### Estimating Model Bias

Using the data-derived parameter maps and the set of microparameters described above, the arterial input function and tissue concentrations for the reference and target regions were forward simulated to generate TACs for every voxel in the brain, creating simulated PET data.

With the simulated PET data fully defined, bias was estimated by comparing data simulated with a two-tissue compartmental model against kinetic modeling analysis performed with a single compartment. By simulating PET data without a challenge, and fitting a model with a challenge, any resultant estimate of ΔBP_ND_ can be attributed solely to bias.

The Multilinear Reference Tissue Model (MRTM2)^13^ was used to measure the BP_ND_ in the striatum (where dopamine receptors are most abundant) with the cerebellum (negligible dopamine D_2_/D_3_ receptor concentration) as the reference tissue and 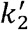 fixed by a first pass fit of MRTM to a high binding region (putamen)^12, 13^. This model was **<u>E</u>**xtended^18^ to E-MRTM2 (**Eq. 4**), accommodating a behavioral challenge using the formulation of Alpert et al.^19^ with a unit step at the onset of the task (t = 40 min) as the activation function. This is similar to the approach taken by Normandin et al.^20^, though with a simpler challenge encoding.

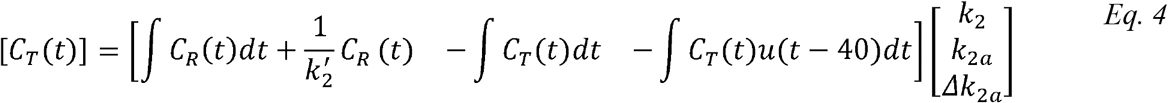

When considering a model with a single tissue compartment, BP_ND_ and ΔBP_ND_ are expressed in terms of a combination of the true efflux rate (k_2_), the apparent efflux rate (k_2a_) and the change in the apparent efflux rate (Δk_2a_)^7^ (**Eqs. Eq**. *5* **&** **Eq**. 6).

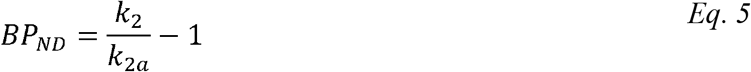

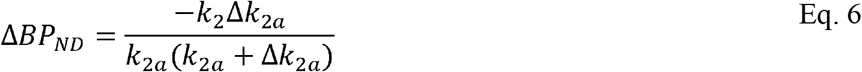

### Mitigating Model Bias in ΔBP_ND_

An analytical approach can be employed to describe the bias that arises from the difference between one-compartment and two-compartment reference tissue models (**Eq. 7**). This difference can be ascribed to a term (E) that depends upon the convolution of the tissue concentration derivative with an exponential function of BP_ND_ and k_4_ ^11^ (**Eq. 8)**.

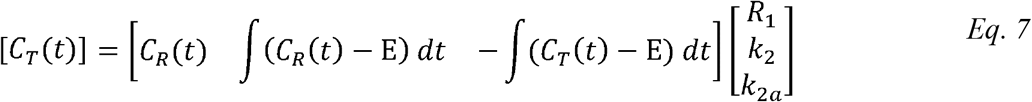

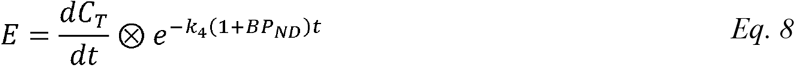

This term is especially pronounced during the uptake period (strong tissue derivative) and in regions with either low binding or low k_4_. Applying a one-compartment tissue model—like MRTM and its variations—assumes that E (and therefore the difference between the two models) is negligibly small. Therefore, to mitigate the bias that arises when it is not, we propose an approach to **<u>D</u>**ebias the contribution of the initial uptake period (DE-MRTM2) where the tissue concentration is changing rapidly. However, to retain kinetic information in the uptake period for estimation of k_2_, we discounted the uptake period for BP_ND_ exclusively by introducing a second “Δk_2a_” term (**Eq. 9)**.

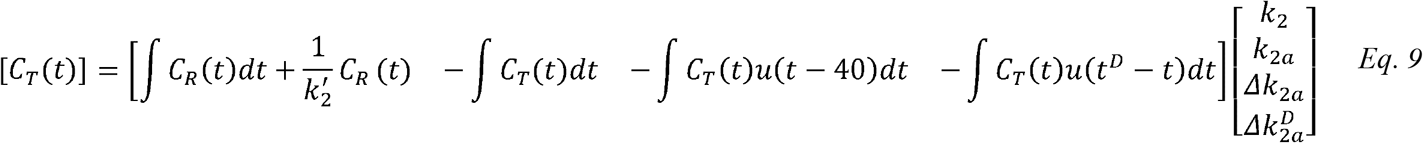

DE-MRTM2 breaks the measurement of *k*_2*a*_ into three periods—initial uptake 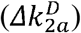, baseline (*k*_2*a*_), task (*Δk*_2*a*_)—while employing the full TAC for estimation of k_2_. The metric of interest, ΔBP_ND_, corresponds to the difference in BP_ND_ between the task and baseline periods. Therefore, **Eqs. Eq**. *5* **&** Eq. 6 still apply for calculating BP_ND_ and ΔBP_ND_ respectively. The additional term 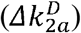 performs its debiasing function by eliminating all contributions of frames prior to the end of the uptake period from the estimation of BP_ND_ and ΔBP_ND_.

To find an appropriate time for ending the uptake period (*t*^*D*^), a series of kinetic analyses were performed, using PET simulations with a k_4_ value within the literature range (0.07 min^-1^)^17^. BP_ND_-dependent TACs were created by sorting voxels within the striatum into bins based on their BP_ND_ values. These TACs were then analyzed using DE-MRTM2 with t^D^ values varied in 3-minute increments consistent with the PET framing of the participant data (21 min, 24 min, 27 min, 30 min, 33 min). Simulated bias (ΔBP_ND_) was plotted against binned BP_ND_ and the t^D^ value that minimized bias was selected for the model.

### Application of DE-MRTM2 to behavioral task data

Measured data from the study participants were utilized to evaluate the impact and performance of DE-MRTM2 relative to conventional analysis. Parametric maps of ΔBP_ND_ were median averaged across participants to reduce the influence of outliers due to motion and other subject-specific effects. Similarly, regional values of ΔBP_ND_ in the gray matter, white matter, and striatum (including subregions—putamen, caudate, nucleus accumbens) were determined using DE-MRTM2 and compared to ΔBP_ND_ values derived from E-MRTM2.

To examine the extent to which this approach to bias mitigation is dependent upon the selection of k_2_’, DE-MRTM2 and E-MRTM2 were fit while the value of k_2_’ was fixed according to one of four estimation methods. In order of highest to lowest estimated value of k_2_’, these methods were:

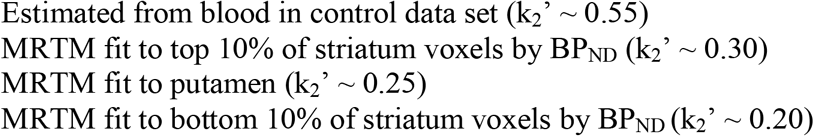

## Results

### Estimating and Mitigating Model Bias in ΔBP_ND_

The value of t^D^ was selected to minimize bias in ΔBP_ND_ across values of BP_ND_ in simulations (**Figure 3**). The absolute minimum of ΔBP_ND_ bias occurred at t^D^ between 27 and 30 minutes post-injection. Therefore, t^D^ was selected to be 27 minutes post-injection to reduce the variance by allowing for one additional frame in estimating baseline BP_ND_.

**Figure 3:**
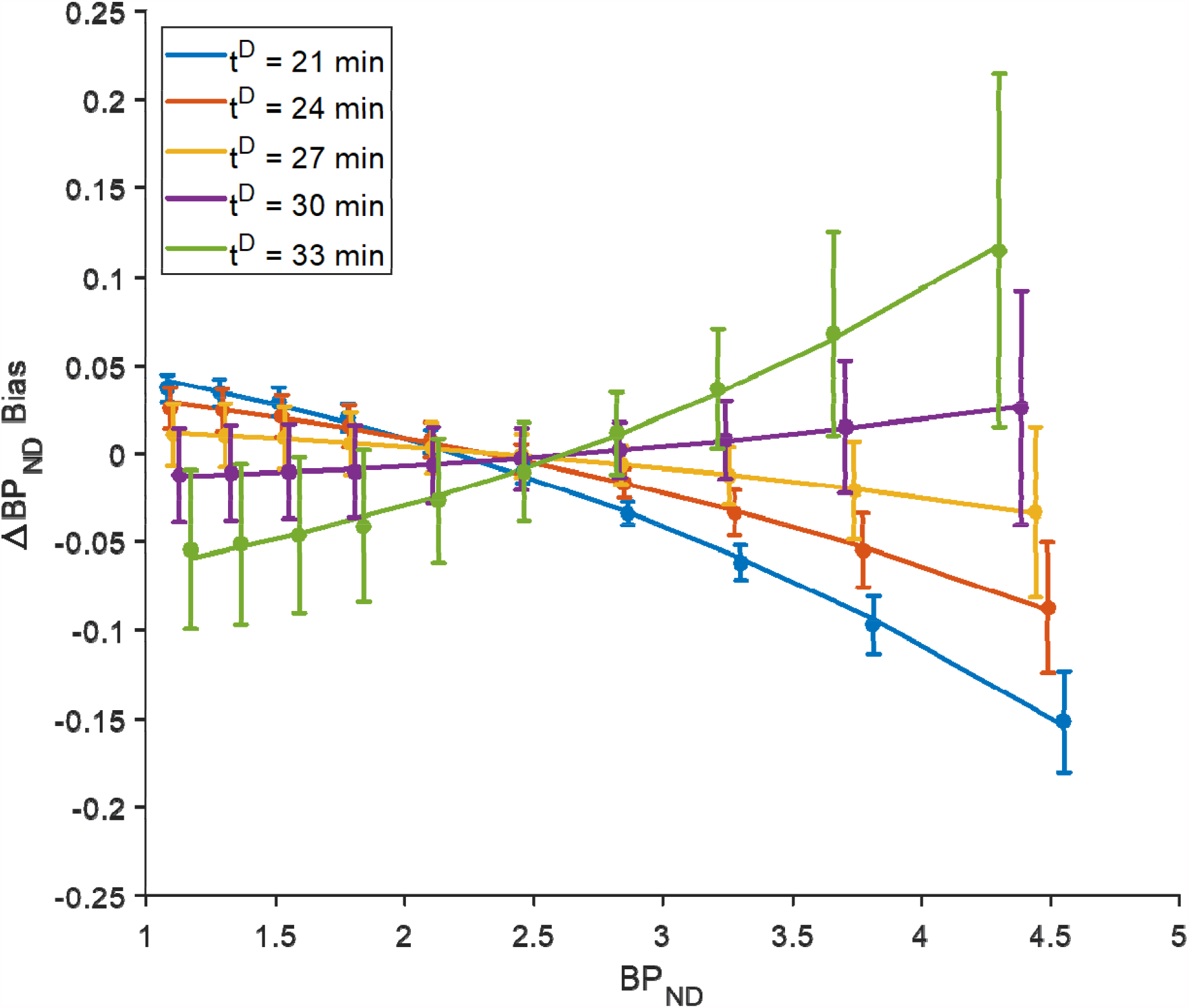
Simulations to determine optimal end of the uptake period (t^D^) for minimal ΔBP_ND_ bias using DE-MRTM2. Error bars indicate standard errors. A value of t^D^ = 27 min was selected for use in analysis.

Model fits of simulated PET data were used to estimate levels of model-induced bias for DE-MRTM2 and E-MRTM2, both at the voxel level (**Figure 4**) and averaged within the nucleus accumbens (**Table 1**). At k_4_ = 0.07, DE-MRTM2 reduced the average bias in the nucleus accumbens from 3.1% to -0.1% and the maximum voxel bias magnitude from 9.8% to 2.4%. At k_4_ = 0.13, DE-MRTM2 reduced the average bias in the nucleus accumbens from 1.2% to -0.3% and the maximum voxel bias magnitude from 3.9% to 2.4%.

**Table 1:** Simulated bias in ΔBP_ND_ using E-MRTM2 and DE-MRTM2 in nucleus accumbens. At lower k_4_, bias becomes more pronounced and DE-MRTM2 becomes more effective, reducing both the average bias and the extent of its range. Absolute ΔBP_ND_ values are provided in parentheses.

|  | E-MRTM2 |  | DE-MRTM2 |  |
| --- | --- | --- | --- | --- |
| $k_4$ (min<sup>-1</sup>) | $\Delta BP_{ND}$ | Range $\Delta BP_{ND}$ | $\Delta BP_{ND}$ | Range $\Delta BP_{ND}$ |
| 0.07 | 0.062 (3.1%) | -2.4% to 9.8%<br>(-0.086 to 0.122) | -0.002 (-0.1%) | -0.6% to 2.4%<br>(-0.020 to 0.018) |
| 0.13 | 0.024 (1.2%) | -0.6% to 3.9%<br>(-0.023 to 0.045) | -0.005 (-0.3%) | -2.4% to 0.4%<br>(-0.021 to 0.012) |

**Figure 4:**
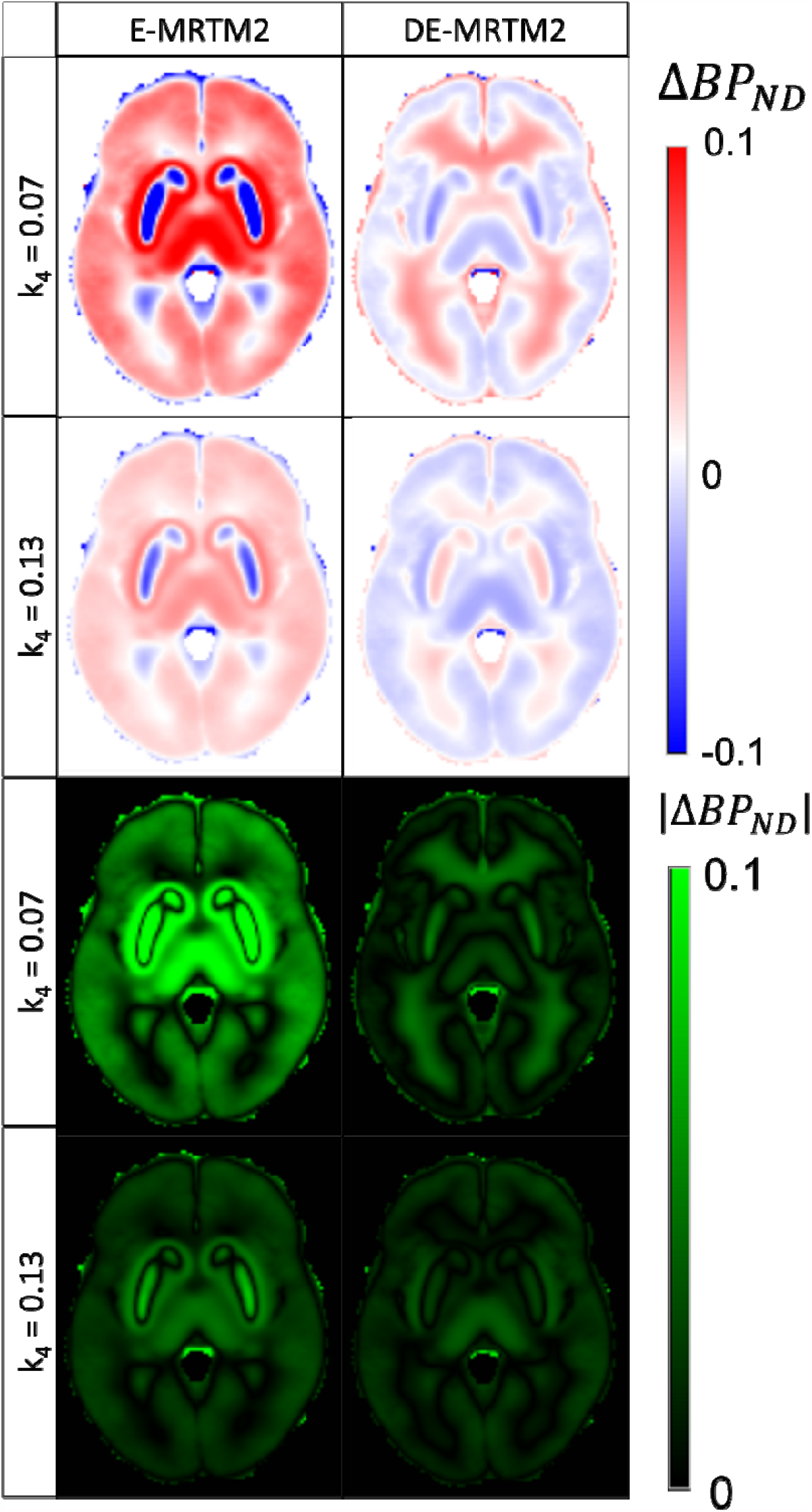
The application of DE-MRTM2 decreases the estimated bias in ΔBP_ND_ in simulations using optimized microparameters and varying k_4._ While the polarity of the remaining bias is dependent on the selection of t^D^ (see **Figure 3**) and the given region, maps of absolute bias demonstrate that DE-MRTM2 reduces the total bias regardless of the underlying value of k_4_.

### Application of DE-MRTM2 to behavioral task data

**Figure 5A** shows the effect of applying DE-MRTM2 to the acquired task-reward data median averaged across participants. While the regional values in the striatum remain similar (**Table 2**), bias in the surrounding areas is mitigated. Specifically, the “halo” of positive values around the striatum becomes less spatially distinct, and positive and negative bias are no longer strongly associated with white and gray matter respectively. **Figure 5B** presents a projection of the ΔBP_ND_ values in **Figure 5A** along the x-axis, highlighting their positioning relative to the boundaries of the striatum ROI.

**Table 2:** Percent change in RAC ΔBP_ND_ before and after mitigating bias in measured participant data. Absolute ΔBP_ND_ values are given in parentheses.

| $\Delta BP_{ND}$ | E-MRTM2 | DE-MRTM2 |
| --- | --- | --- |
| Striatum | -2.7% (-0.07) | -2.2% (-0.06) |
| White Matter | 6.1% (0.02) | 2.5% (0.01) |
| Gray Matter | -4.3% (-0.01) | 6.6% (0.01) |
| Striatal Subregions | E-MRTM2 | DE-MRTM2 |
| Putamen | -1.9% (-0.06) | -1.9% (-0.06) |
| Caudate | -4.5% (-0.10) | -3.1% (-0.07) |
| Nucleus Accumbens | -1.5% (-0.03) | -1.8% (-0.04) |

**Figure 5:**
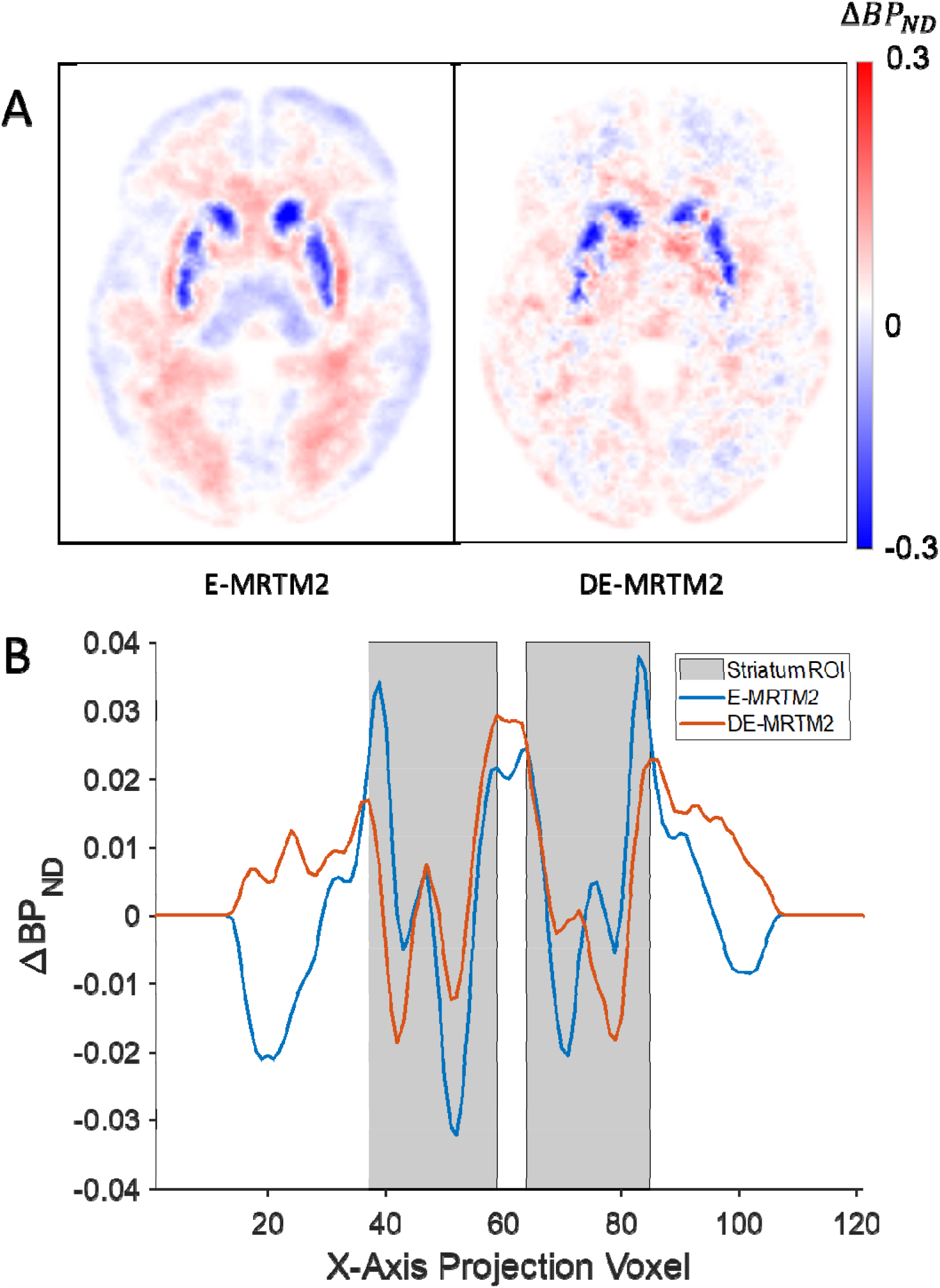
**(A)** Change in non-displaceable binding potential in response to behavioral task. Voxel maps are median averaged across all 69 study participants. The map created using E-MRTM2 on the left includes both the effects of task response and model-based bias. The DE-MRTM2 map on the right removes much of the model bias. **(B)** 1D projection of ΔBP_ND_ maps onto the x-axis. The volume was first projected into the coronal plane (2D), before being projected onto the x-axis (1D). The projection was limited to the axial and coronal planes that contain the striatum.

**Figure 6** shows the relationship between DE-MRTM2 and the selection of k_2_’. The E-MRTM2 fits are highly dependent on the value of k_2_’ fixed prior to analysis. The conventional method for selecting the value of k_2_’ (MRTM fit to a high binding region like putamen) produces the least biased ΔBP_ND_ maps, while setting the value of k_2_’ either too high or too low causes associated overestimation and underestimation of ΔBP_ND_ respectively. Using DE-MRTM2 however, the bias is reduced and becomes far less dependent on the selection of k_2_’.

**Figure 6:**
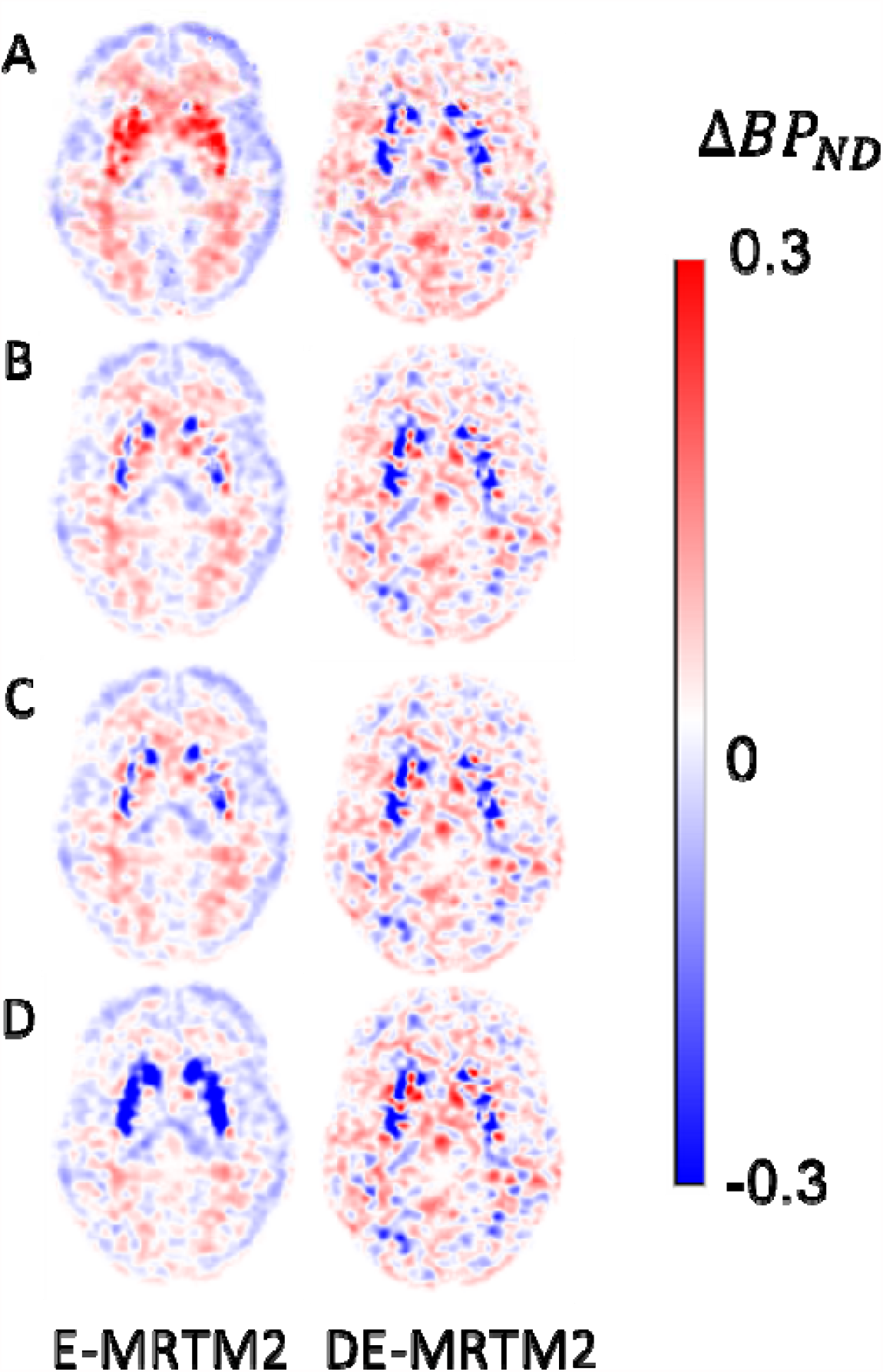
Effect of debiasing at different levels of k_2_’. (A) k_2_’ = 0.55, estimated from blood plasma fits in control data set. (B) k_2_’ = 0.30, estimated from MRTM fits of top 10% of striatum voxels by BP_ND_. (C) k_2_’ = 0.25, estimated from MRTM fit to regional putamen TAC. (D) k_2_’ = 0.20, estimated from MRTM fits of bottom 10% of striatum voxels by BP_ND_.

## Discussion

We investigated model-based bias in the estimation of change in ^11^C-raclopride binding potential in response to a behavioral reward task experiment. Simulations were designed to reflect the conditions of the acquired data, with both baseline and task conditions performed in a single imaging session. A method for selectively discounting the initial uptake period in extended-MRTM2 estimation of BP_ND_ and ΔBP_ND_ was proposed to decrease model bias associated with the use of a one-tissue compartment model. Simulated levels of bias in ΔBP_ND_, which vary spatially with BP_ND_, were also observed to depend on k_4_ (**Table 1**). In the human task data, the average measured ΔBP_ND_ in striatum changed from -2.7% of BP_ND_ to -2.2% of BP_ND_ after debiasing. DE-MRTM2 was also shown to greatly decrease the dependence of bias in ΔBP_ND_ upon k_2_’.

The primary motivation for minimizing sources of bias in ΔBP_ND_ is that they may confound or obscure true experimental results of interest. Therefore, when evaluating the impact of an approach for reducing model bias, it is important to consider brain regions of differing experimental relevance separately. In both the parametric maps and regional averages of ΔBP_ND_, the low-binding extrastriatal regions exhibited the greatest reduction in bias after applying DE-MRTM2. However, the relatively low affinity of RAC for dopamine D_2_/D_3_ receptors (K_d_ = 1.3 nM)^21^ makes it an ineffective choice for extrastriatal measurements relative to a higher affinity tracer like ^18^F-fallypride^22^. Recent work has further indicated that RAC PET quantified using the cerebellum as a reference region overestimates specific binding in cortex and is therefore unsuited to measuring extrastriatal dopamine binding, much less its transient changes^21^. Therefore, any efforts to quantify extrastriatal dopamine responses using RAC should be mindful not only of the sources of model bias that may be present, but also the challenges and limitations of the approach overall.

Within the striatum, the modest impact of DE-MRTM2 implies that there may be a relatively high underlying value of k_4_ for RAC. In the control data set^16^, two-tissue compartmental model fits of the putamen data provided k_4_ estimates of approximately 0.13 min^-1^, which was on the higher end of the examined range and produced lower levels of bias in simulations. If the reward data set features similar k_4_ values, the overall value of the error term, E (**Eq. 8**) would be small at high values of BP_ND_, implying low model bias, particularly relative to the genuine task response signal in the striatal subregions where it is the strongest.

In the experimental results, each of the striatal subregions responded to debiasing differently (**Table 2)**. The use of debiasing reduced average apparent displacement in caudate, increased apparent displacement in nucleus accumbens, and had little effect in putamen. These differential bias effects are likely related to precisely where the borders of each anatomically-derived ROI label fall with respect to the spatial gradients in binding potential. Using E-MRTM2, striatal regions with high BP_ND_ tend to overestimate challenge response, while striatal regions with low BP_ND_ tend to underestimate it, forming a “halo” of apparent task-related increases in BP_ND_ around the edges of the striatum^11^. Applying DE-MRTM2, the parametric map of ΔBP_ND_ is more tightly matched to the boundaries of the regional labels. This can be observed in **Figure 5B** where using E-MRTM2, the bias halo (positive peaks) in the projection of ΔBP_ND_ falls within the striatum. With DE-MRTM2, these bias peaks decrease in magnitude and coincide more closely with the edges of the striatum ROI. In ROI analysis, voxels with differing bias polarities within the same region work in opposition, causing the extent of the regionally defined bias to be understated. Given the gradients of bias within regions, literature reports based upon ROI analyses will be sensitive to the details of regional delineation.

In addition to the model bias that arises when simplifying from 4-parameter FRTM to 3-parameter MRTM, further bias is introduced when reducing to 2-parameter MRTM2^12^. This reduction is necessary because 3-parameter MRTM fits of ΔBP_ND_ yield far noisier parametric maps and corresponding increases in the variance of regional fits. In our previous work, we observed that fixing k_4_ to the optimal value resulted in both a decoupling of the correlation between BP_ND_ and k_2_’ and reduced bias in estimations of BP_ND_^11^. By fitting estimates of BP_ND_ and ΔBP_ND_ only after the end of the initial uptake period (t^D^), we were able to mitigate the bias introduced by the selection of k_2_’ even when using a single-compartment model.

Our study had several limitations. Because the experimental protocol was designed to investigate group differences in dopamine response based on task performance—thereby encoding a learning behavior—all participants performed the rewarded task^23^. Pure baseline acquisitions (without task) were not acquired. Had it been available, such control data—ideally with the same participants—could have been used to better distinguish task response from other confounding influences like model bias. It could also be illuminating to have a cohort whose task experiment commenced later in the scan (e.g. 60-minutes post-injection). As all data prior to time t^D^ was excluded from the fit of BP_ND_ and its task response, there was only a window of 13 minutes for the estimation of baseline BP_ND_ compared to the 50 minutes remaining in the scan for the estimation of ΔBP_ND_. Preliminary simulations (not shown) have indicated that later task start times may reduce the underestimation of genuine task response effects, making this a promising avenue for future work.

Finally, these results are limited to RAC and specifically to the context of an intrascan behavioral task challenge paradigm. Pharmacological challenges administered within-scan produce larger responses and may be amenable to more complex models capable of observing greater temporal detail in the responses^3^. Models fit to data acquired using other tracers that exhibit different kinetic properties may also be impacted by bias differently, especially if the values of k_4_ and k_2_’ are different, or if different assumptions are violated. Tracers that exhibit more non-specific binding, receptor internalization, specific binding in the reference region, or signal attributable to blood volume may be more biased by those sources^10^ than by the discrepancy between one compartment and two compartment configurations.

In conclusion, the impact of bias arising from fitting a model with a 1TCM configuration to two-compartment data has been investigated in the context of an intrascan behavioral task challenge paradigm using RAC PET. Computer simulation studies demonstrated that for RAC, model bias can contribute to the overall observed effect while still being smaller than the total response reported in this particular behavioral study. Selectively discounting the initial uptake period in the estimation of BP_ND_ and ΔBP_ND_ can help reduce this model-based bias, while maintaining the low variance and high stability of a two-parameter model.

## Data Availability

For inquiries into data availability, please contact the corresponding author.

## Acknowledgements

We would like to thank Rajesh Narendran for making available to us a set of previously published RAC data with arterial blood sampling^16^ for comparison.

This work was partly supported by National Institute of Biomedical Imaging and Bioengineering Grant 5R01EB014894-02, National Institute of Mental Health Grant Number R01MH080243, National Institute of Neurological Disorders and Stroke Grant Number R01NS112295, National Institute of General Medical Sciences Grant T32 GM008313, NIH Blueprint for Research Science Grant T90DA022759/R90DA023427, and NIH Shared Instrumentation Grant S10RR023043.

